# Do no harm - re-evaluating the risks of overtreatment in community-wide tuberculosis screening

**DOI:** 10.1101/2025.09.18.25335917

**Authors:** Rein M.G.J. Houben, Lara D. Veeken, Alvaro Schwalb, Daniel J. Grint, Sean Wasserman, Reinout van Crevel, Katherine C. Horton

## Abstract

**Background:** Community-wide screening is a crucial strategy to end tuberculosis (TB), but a common concern is potential harm from overtreatment following false positive diagnoses. However, current reference standards determining test performance have limitations, with implications for prevalence thresholds and treatment decisions for community-wide screening.

**Methods:** We estimated coverage of community-wide screening at a prevalence threshold of 0.5% (current global standard), 0.25%, and 0.1% for adult pulmonary TB. We considered test performance for Xpert Ultra against different reference standards (sputum culture, plus clinical evaluation, plus disease progression within two years). Potential harm was estimated through disability adjusted life years (DALYs) incurred or averted by treatment. We report net specificity, positive predictive value (PPV), the ratio of false positives to true positives, and DALYs averted for (non-)treatment based on different reference standards.

**Results:** A lower threshold would increase screening coverage from the current 42% to 84% (0.25% threshold) and 89% (0.1% threshold) of the global TB burden. In a population of 100,000 with 0.5% prevalence, specificity was 99.5% for community screening, but increased to 99.7% using disease progression as reference standard, with PPV increasing from 45 to 66%. In addition, estimated harm of withholding appropriate treatment was approximately 1,200 times higher compared to providing inappropriate treatment, with treatment initiation after a positive Xpert Ultra increasing overall DALYs averted (median 5,977 versus 3,750).

**Discussion:** The benefit of TB treatment following a positive molecular test in community-wide screening likely outweighs the harm associated with possible overtreatment, supporting expanding coverage of simplified community-wide screening.

**Brief summary:** A concern with community-wide tuberculosis screening is the potential for overtreatment. We evaluated diagnostic reference standards and relative health costs of (non-)treatment, finding that after a positive molecular test, the benefit of initiating tuberculosis treatment likely outweighs its harm.

## Introduction

An estimated 10 million people develop tuberculosis (TB) each year, resulting in 1.3 million deaths (1). While community-wide screening contributed to strong historical declines (2–6) and has shown potential in modern times (7), it is currently underutilised. (2) Increasing coverage of community screening has many challenges (2), including the current policy threshold of 0.5% prevalence of bacteriologically confirmed TB. A key concern is the potential harm from overtreatment driven by increasingly high numbers of false positive diagnoses and lower positive predictive values (PPVs), as a diagnostic algorithm with fixed specificity is applied in populations with low(er) prevalences. (8)

However, recent ACF studies have not observed extensive overtreatment (9), even when conducted in populations with prevalence well below the current threshold of 0.5%. (7) Part of the explanation is that specificity is not fixed, but instead increases as prevalence drops, a widely recognised phenomenon in diagnostics (10), and confirmed for TB in a recent analysis of PCR test performance. (11) An important driver of changing specificity is the likely (more) rapid change in performance of the reference standard test against which the performance of the diagnostic is measured as prevalence falls. (10)

In the TB response, sputum culture has been the reference standard test for decades, even after PCR-based tests became widely used. (12,13) The assumption is that if *Mycobacterium tuberculosis* (*Mtb*) can be grown from an individual’s respiratory sample, the disease process or symptoms under investigation are due to *Mtb*. In turn, this means the individual will benefit from taking TB drugs, outweighing the harm from receiving treatment (through e.g. adverse events or costs). However, the reverse is also implicitly assumed: a negative culture argues against treatment, especially in the absence of typical TB symptoms or chest X-ray abnormalities.

However, sputum culture is imperfect, as illustrated by the fact that with repeated cultures more individuals test positive at least once (14,15). Inherently, this implies false negative culture results do occur, likely due to factors such as bacterial death or contamination between sample collection and inoculation. (16) Such false negative results will likely increase with community screening due to a higher proportion of paucibacillary samples (17) and increased distance between point of collection and culture-processing facility.

Sputum culture is also an imperfect predictor of who would benefit from treatment. Follow-up of untreated bacteriologically-negative individuals with pathology consistent with TB has shown a high proportion will start treatment within two years of negative test results. (18–20) Also, historically impactful community-wide screening efforts often included treatment of many (50% or more) individuals without bacteriological, let alone culture, confirmation. (4,6)

With culture as an imperfect reference standard, false negative results are incorrectly counted as false positive results for the comparator test (e.g. PCR-based tools under evaluation (21)). As a consequence, we are overestimating the risk of overtreatment and harm in low(er) prevalence communities, when in fact treatment based on such tests could be beneficial to the individual.

For community-wide screening to have meaningful impact, we need to increase coverage and reduce the human and financial resources required. If the performance of PCR-based bacteriological tests is better than currently assumed, the pathway from initiating screening to treatment initiation could be simplified by dropping confirmatory sputum culture or clinical examination, and immediately start TB treatment. (2)

Here, we evaluate the issue of overtreatment in the context of community-wide screening and shifting reference standards, balancing the risk for doing harm versus the potential benefit of treating those with a positive PCR test for the individual and their community.

## Methods

### Prevalence threshold and coverage of community screening

To estimate the proportion of global prevalent TB that is covered by the current threshold of 0.5% for community screening, we converted 2023 WHO country-level incidence estimates to an approximate prevalence of adult TB. (22) A simple national-level prevalence-to-incidence ratio was applied for each country, using the 2000-2014 WHO estimates for prevalence and incidence. Detailed methods are in supplementary material 1. We compared our estimates with reported prevalence from national TB prevalence surveys, and calculated the proportion of prevalent TB burden covered by prevalence thresholds of 0.5% (current threshold), 0.25%, and 0.1% using the country-level prevalence as the metric.

### Test characteristics by reference standard

To explore the impact of different reference standards on test performance, we investigated the performance of Xpert Ultra, a widely used PCR-based sputum test, in community-wide screening across reference standards and population prevalences.

We considered a population of 100,000 adults with a 0.5% prevalence of sputum culture-confirmed pulmonary TB. Performance of sputum Xpert Ultra against sputum culture in community-wide screening was based on the trend in a recent evaluation of specificity in community settings (11,23,24), with a specificity of 99.5% and a pooled sensitivity of 82.5% among individuals screened positive on symptoms and/or abnormal chest X-ray in prevalence surveys (see supplementary material 2 for derivation). (11)

Based on those metrics, individuals were distributed between true positive (TP), false positive (FP), true negative (TN), and false negative (FN), using Xpert-positive/culture-positive, Xpert-positive/culture-negative, Xpert-negative/culture-negative and Xpert-negative/culture-positive, respectively. From these results, we calculated the FP:TP ratio, population prevalence ((TP+FN)/(TP+FP+TN+FN)), positive predictive value (PPV, calculated as TP/(TP+FP)) and negative predictive value (NPV, calculated as TN/(TN+FN)), number needed to screen (NNS), and number needed to harm (NNH).

For the impact of changing the reference standard from the standard of sputum culture, we used results from a recent community-wide screening trial in Uganda, which followed up individuals with an initial positive Xpert Ultra trace result. (20)In brief, individuals with a trace-positive Xpert received a detailed baseline evaluation including symptom survey, a second Xpert Ultra, two sets of sputum liquid and solid cultures, lipoarabinomannan test if HIV positive, chest X-ray, and chest CT scan (see supplementary material 2 for details). This approach identified 18% of trace-positive/culture-negative (i.e. counted as FP in baseline scenario) as requiring treatment, 50% of which were based on a positive repeat non-culture bacteriological test. Untreated individuals were followed up actively for up to two years, during which 25% of the remainder were referred for treatment, mostly within six months (Supplementary Table 5).

Using these data, we constructed two alternative reference standards: 1) culture for all + evaluation among Xpert Ultra positive individuals, and 2) culture for all + evaluation among Xpert Ultra positive individuals + two-year follow-up among Xpert Ultra positive individuals without treatment recommendation at baseline. We assumed that the additional treatment initiations indicated false negative culture results, and instead the initial Xpert result was correct. To reflect this, we reassigned individuals from FP to TP in line with the 18% found positive at initial evaluation, and further 25% for the two year follow-up, and then recalculated the other outcomes.

Calculations were then repeated with a population with a TB prevalence of 0.25% and 0.1%, where the initial specificity of Xpert Ultra was adjusted by extending observed trends to lower prevalences. (23)

### Benefit versus harm

To quantify the relative health benefit and harm of treatment based on the PCR test, we considered the disability adjusted life years (DALYs) averted by treating someone after a TP diagnosis and incurred from treating someone after a FP diagnosis).

Estimates for DALYs averted considered both the individual (period of ill health, risk of death, and post-TB sequelae) and community (secondary episodes following *Mtb* transmission). We extracted estimates from the literature and published models. (25–27) For the main analysis, we reduced the mid-point value by 50% to reflect that not all negative health effects (e.g. transmission, lung damage leading to post-TB disease) would be averted by earlier diagnosis and treatment, and to generate a conservative estimate of benefit. See Table 2 and Supplementary Table 10 for details.

Estimates of DALYs incurred due to TB treatment included serious adverse events (SAE), such as hepatoxicity, as well as tolerability issues like daily vomiting (26). In the absence of published values, we generated estimates by using reported rates of SAE events as a lower bound (28)) and tolerability (e.g. repeated vomiting, in ~10% of individuals (26)), and assigned inflated utility costs (0.35 for SAE, 0.2 for intolerance). (29) Finally, we set the average duration at 0.1 year (~5 weeks), which is approximately double the estimated duration of hepatoxicity, the most common SAE. (28) In absence of data or conversion, we did not include DALYs for the financial impact of treatment, including catastrophic costs due to patient costs incurred during treatment (for both TP and FP), or pre-treatment (for TP and FN). See table 2 and Supplementary Table 10 for details.

We estimated the ratio between DALY averted (for treating TP) and incurred (for treating FP). In addition, we estimated total DALYs, which were assigned for untreated TB (counted as incurred for FN) and for TB treatment (counted as incurred for TP and FP). Total averted DALYs were calculated for each reference standard, as well as the incremental averted DALYs for the alternative reference standards (see Supplementary Table 11 for details).

### Sensitivity analyses

We repeated analyses with baseline sensitivity of Xpert for 70%, 60%, and 50% to explore the robustness of results to the assumed sensitivity of Xpert Ultra when applied in the community, and align with the range of sensitivity explored in the WHO target product profiles for tuberculosis screening tests. (30) These were estimates in the absence of strong data on sensitivity in the full prevalence range.

## Results

### Prevalence threshold and coverage of community screening

The average prevalence-to-incidence ratio in WHO data was 1.35:1 over the 2000-2014 period (Supplementary Figure 1), and provided 2023 prevalence estimates of adult bacteriologically confirmed TB (Supplementary Table 2). Supplementary Figure 2 compares our 2023 estimates with the same metric from prevalence surveys conducted between 2010 and 2020, which showed strong overlap in almost all countries.

**Figure 1:**
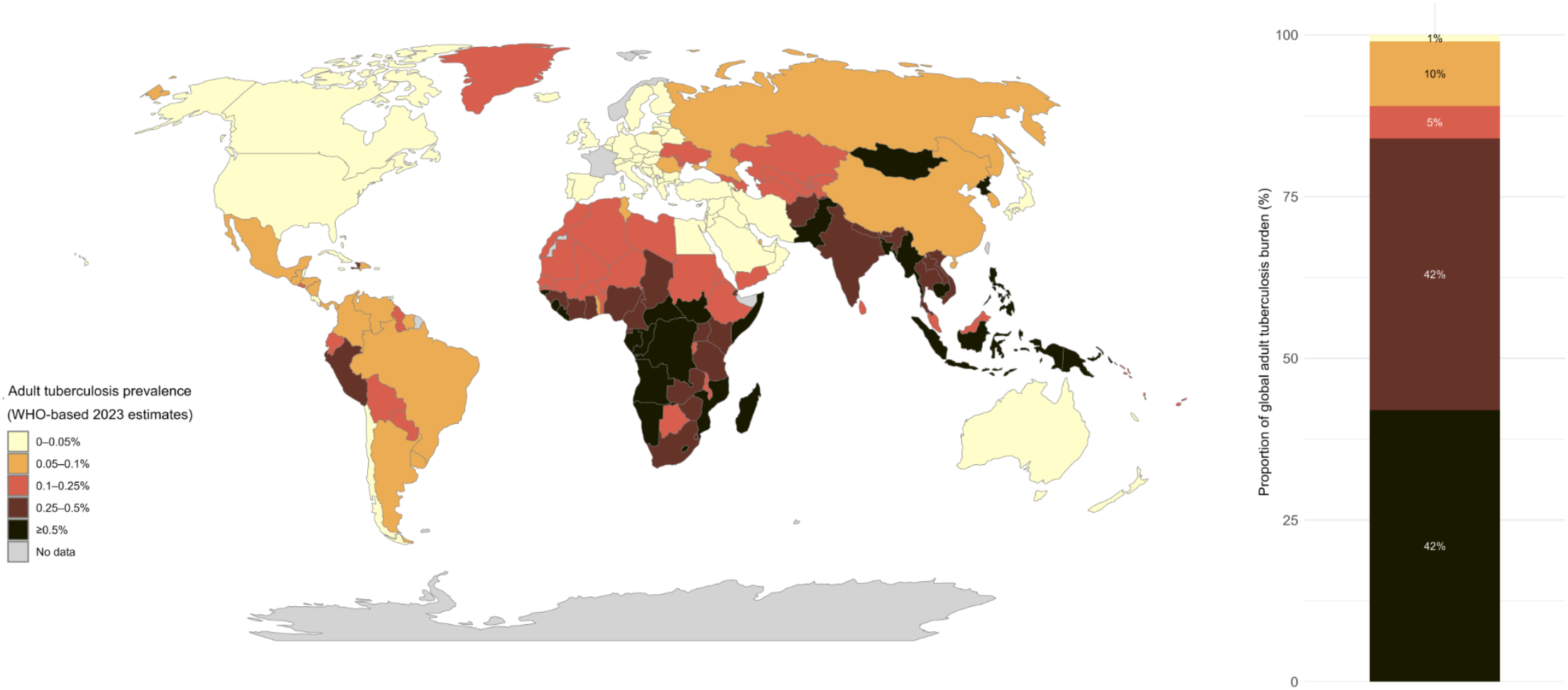
Proportion of adult tuberculosis prevalence (by threshold) Figure shows countries that could initiate community-wide screening among adults at different prevalence thresholds (map, left). Stacked bar chart shows the proportion of all estimated prevalent adult TB that would be covered by different thresholds.

**Figure 2:**
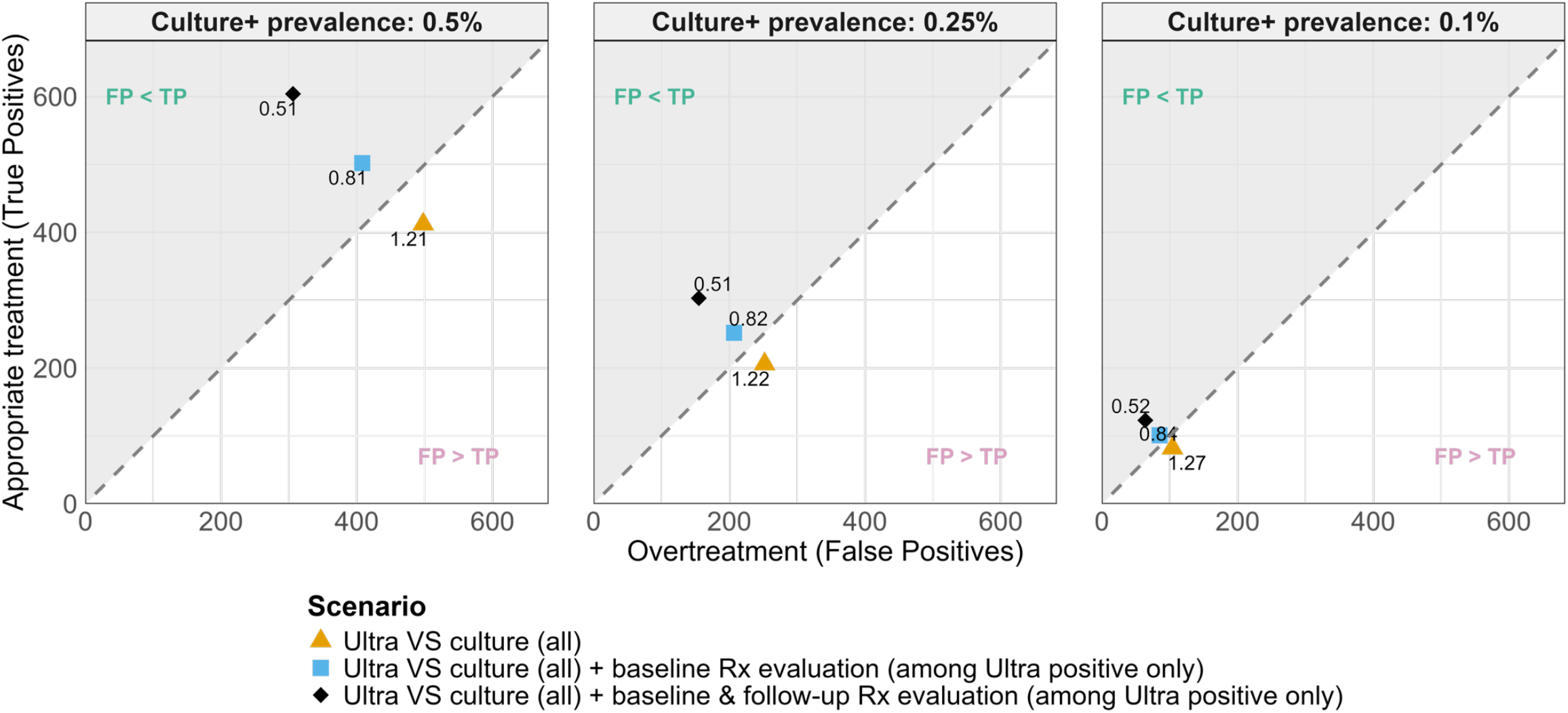
Balance between overtreatment and appropriate treatment, among a hypothetical population of 100,000 individuals. Figures show the balance between the number of individuals receiving overtreatment (i.e. treatment after a false positive test, x-axis) and appropriate treatment (after a true positive test, y-axis), under different reference standards (markers) and prevalence of sputum culture positive TB (plots). Diagonal line indicates 1:1 false-to-true positive ratio. Base sensitivity and specificity are 82.5% (all plots) and 99.5 (left plot), 99.7 (middle plot) and 99.9% (right plot).

Figure 1 shows the proportion of TB covered by the various thresholds, highlighting that coverage would increase from 42% to 84% and 89% of the TB burden if the threshold were moved to 0.25% and 0.1%, respectively.

### Test performance

Further baseline evaluation or follow-up changed the classification of Xpert-positive results not supported by culture. Table 1 shows the test performance characteristics with a changing reference standard. With a population of 100,000 and a culture-confirmed TB prevalence of 0.5%, the baseline sensitivity (82.5%) and specificity (99.5%) based on a sputum culture reference standard would result in 412 TP diagnoses for 498 FP diagnoses (FP:TP ratio of 1.20:1), and a PPV of 0.45. Assuming further baseline evaluation among Xpert Ultra-positive individuals would correct FN culture results, with 90 FP Xpert results reclassified as TP. In terms of test performance, the specificity of Xpert increases to 99.6%. In addition, the population prevalence increases to 0.59%. Sensitivity of Xpert Ultra against this endpoint is increased to 85.1%, with a PPV of 0.55 and a more favourable FP:TP ratio of 0.81:1. When the reference standard also includes progression within two years, a further 102 individuals move from FP to TP, leading to a specificity, prevalence of disease, and sensitivity of 99.7%, 0.69%, and 87.3%, respectively. The PPV and FP:TP improve to 0.66 and 0.51:1, respectively.

**Table 1:**
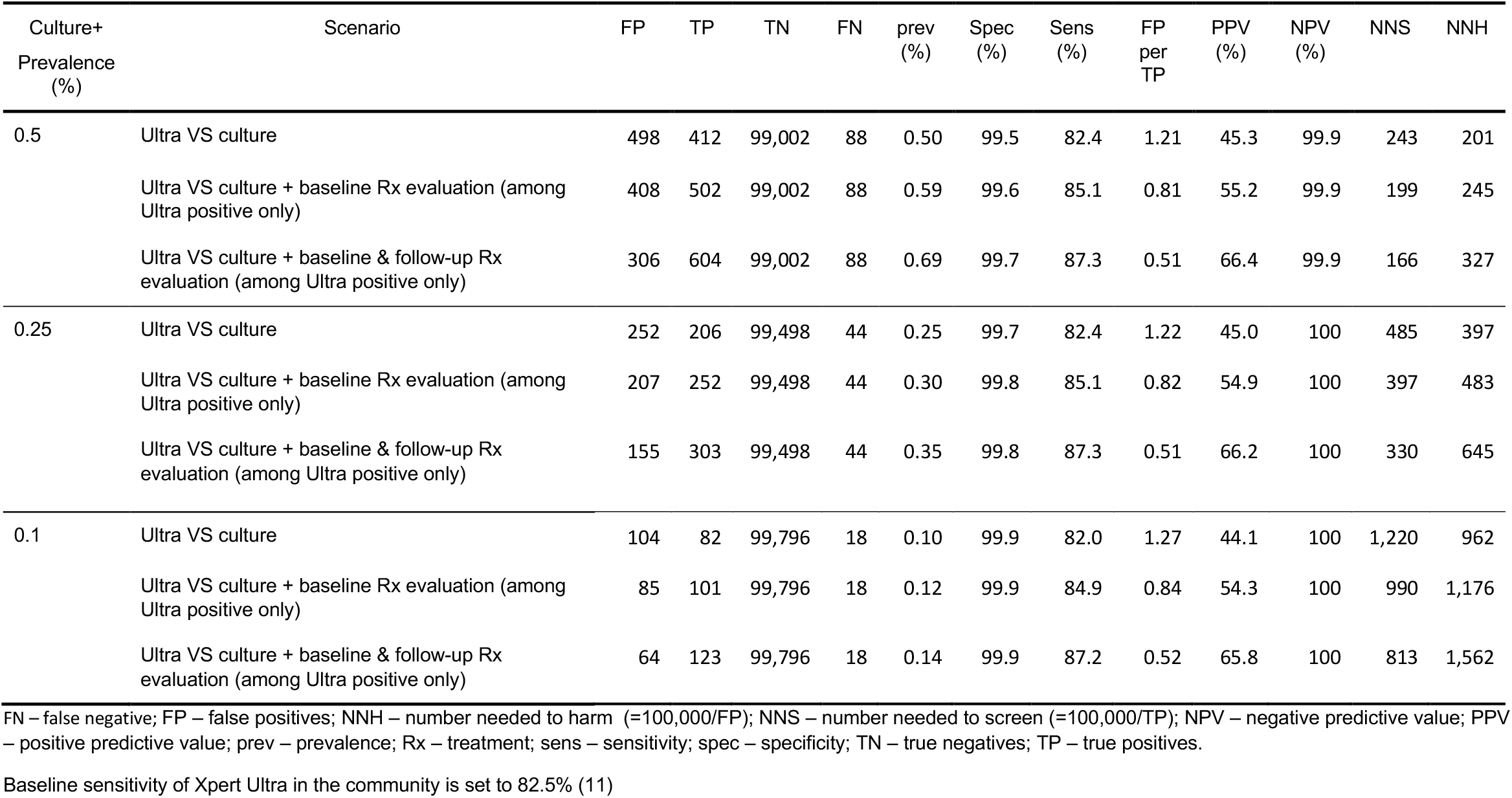
Screening test performance with extended reference standard, among a hypothetical population of 100,000 individuals.

| Culture+<br>Prevalence<br>(%) | Scenario | FP | TP | TN | FN | prev<br>(%) | Spec<br>(%) | Sens<br>(%) | FP<br>per<br>TP | PPV<br>(%) | NPV<br>(%) | NNS | NNH |
| --- | --- | --- | --- | --- | --- | --- | --- | --- | --- | --- | --- | --- | --- |
| 0.5 | Ultra VS culture | 498 | 412 | 99,002 | 88 | 0.50 | 99.5 | 82.4 | 1.21 | 45.3 | 99.9 | 243 | 201 |
|  | Ultra VS culture + baseline Rx evaluation (among Ultra positive only) | 408 | 502 | 99,002 | 88 | 0.59 | 99.6 | 85.1 | 0.81 | 55.2 | 99.9 | 199 | 245 |
|  | Ultra VS culture + baseline & follow-up Rx evaluation (among Ultra positive only) | 306 | 604 | 99,002 | 88 | 0.69 | 99.7 | 87.3 | 0.51 | 66.4 | 99.9 | 166 | 327 |
| 0.25 | Ultra VS culture | 252 | 206 | 99,498 | 44 | 0.25 | 99.7 | 82.4 | 1.22 | 45.0 | 100 | 485 | 397 |
|  | Ultra VS culture + baseline Rx evaluation (among Ultra positive only) | 207 | 252 | 99,498 | 44 | 0.30 | 99.8 | 85.1 | 0.82 | 54.9 | 100 | 397 | 483 |
|  | Ultra VS culture + baseline & follow-up Rx evaluation (among Ultra positive only) | 155 | 303 | 99,498 | 44 | 0.35 | 99.8 | 87.3 | 0.51 | 66.2 | 100 | 330 | 645 |
| 0.1 | Ultra VS culture | 104 | 82 | 99,796 | 18 | 0.10 | 99.9 | 82.0 | 1.27 | 44.1 | 100 | 1,220 | 962 |
|  | Ultra VS culture + baseline Rx evaluation (among Ultra positive only) | 85 | 101 | 99,796 | 18 | 0.12 | 99.9 | 84.9 | 0.84 | 54.3 | 100 | 990 | 1,176 |
|  | Ultra VS culture + baseline & follow-up Rx evaluation (among Ultra positive only) | 64 | 123 | 99,796 | 18 | 0.14 | 99.9 | 87.2 | 0.52 | 65.8 | 100 | 813 | 1,562 |

FN – false negative; FP – false positives; NNH – number needed to harm (=100,000/FP); NNS – number needed to screen (=100,000/TP); NPV – negative predictive value; PPV – positive predictive value; prev – prevalence; Rx – treatment; sens – sensitivity; spec – specificity; TN – true negatives; TP – true positives.
Baseline sensitivity of Xpert Ultra in the community is set to 82.5% (11)

Diagnostic reclassification would also change the balance between benefit and potential harm of treating Xpert-positive culture-negative individuals. Table 1 shows how the NNS and NNH shift with the reference standard. Against the two-year follow-up, the NNS dropped from 243 to 166, and NNH increased from 201 to 327. **Figure 2** shows the balance between the number of individuals receiving appropriate treatment (i.e. TP) and those who are overtreated (i.e. FP), and how this balance shifts in favour of treating solely based on initial Xpert result, depending on the reference standard used.

We find similar trends in changed true prevalence and ratio of appropriate treatment versus overtreatment (Table 1, Figure 2) between reference standards when considering culture-confirmed prevalences of 250 and 100/100,000, where baseline specificities are 99.7% and 99.9%, respectively.

### Benefit versus harm

Combining each component in Table 2, we estimated that the benefit of appropriate treatment for TB was 11.6 DALYs averted (range 2.8-23.6), and the cost of undergoing treatment was 0.009 DALY incurred (range 0.005-0.046) (Supplementary Figure 6). A point value ratio of over 1,200:1 suggests a substantially larger benefit from treating one additional individual appropriately compared to the harm from overtreating one individual (Figure 3).

**Table 2:**
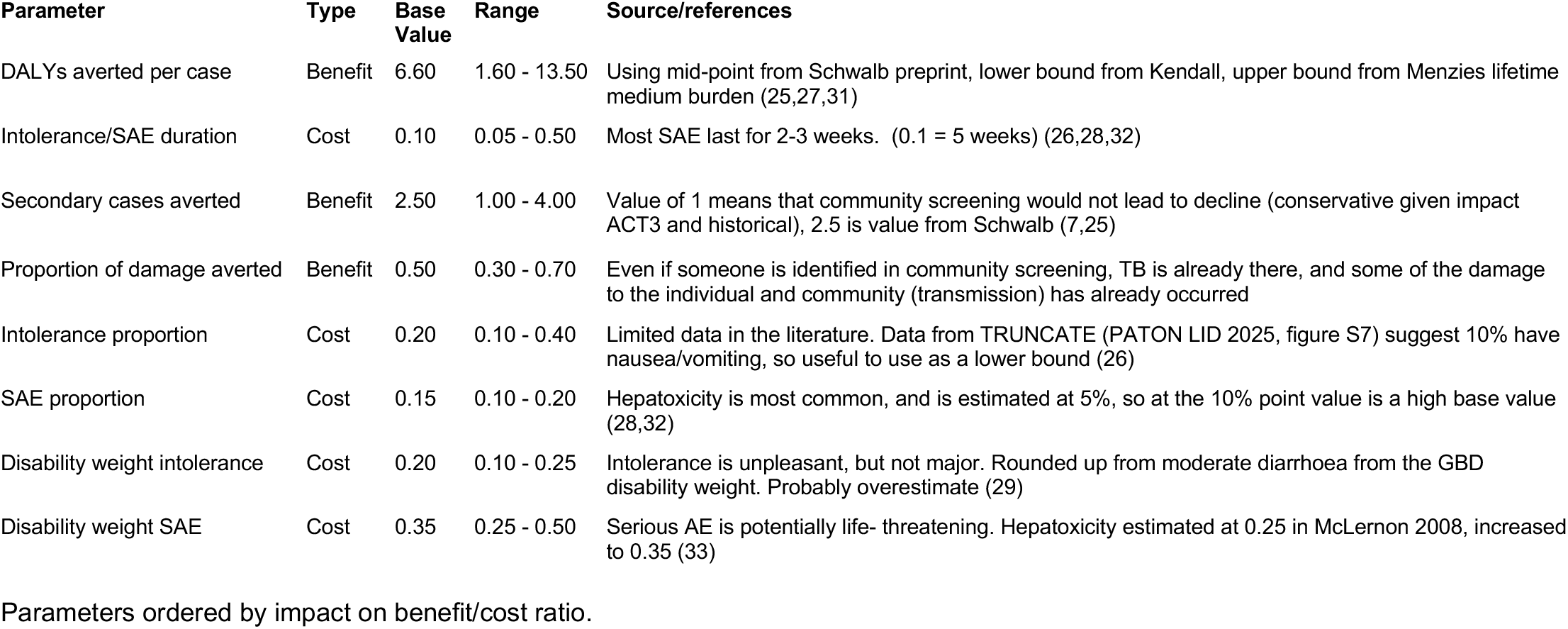
Parameters for estimating benefit and harm of treatment.

| Parameter | Type | Base Value | Range | Source/references |
| --- | --- | --- | --- | --- |
| DALYs averted per case | Benefit | 6.60 | 1.60 - 13.50 | Using mid-point from Schwalb preprint, lower bound from Kendall, upper bound from Menzies lifetime medium burden (25,27,31) |
| Intolerance/SAE duration | Cost | 0.10 | 0.05 - 0.50 | Most SAE last for 2-3 weeks. (0.1 = 5 weeks) (26,28,32) |
| Secondary cases averted | Benefit | 2.50 | 1.00 - 4.00 | Value of 1 means that community screening would not lead to decline (conservative given impact ACT3 and historical), 2.5 is value from Schwalb (7,25) |
| Proportion of damage averted | Benefit | 0.50 | 0.30 - 0.70 | Even if someone is identified in community screening, TB is already there, and some of the damage to the individual and community (transmission) has already occurred |
| Intolerance proportion | Cost | 0.20 | 0.10 - 0.40 | Limited data in the literature. Data from TRUNCATE (PATON LID 2025, figure S7) suggest 10% have nausea/vomiting, so useful to use as a lower bound (26) |
| SAE proportion | Cost | 0.15 | 0.10 - 0.20 | Hepatotoxicity is most common, and is estimated at 5%, so at the 10% point value is a high base value (28,32) |
| Disability weight intolerance | Cost | 0.20 | 0.10 - 0.25 | Intolerance is unpleasant, but not major. Rounded up from moderate diarrhoea from the GBD disability weight. Probably overestimate (29) |
| Disability weight SAE | Cost | 0.35 | 0.25 - 0.50 | Serious AE is potentially life- threatening. Hepatotoxicity estimated at 0.25 in McLernon 2008, increased to 0.35 (33) |

Parameters ordered by impact on benefit/cost ratio.

**Figure 3:**
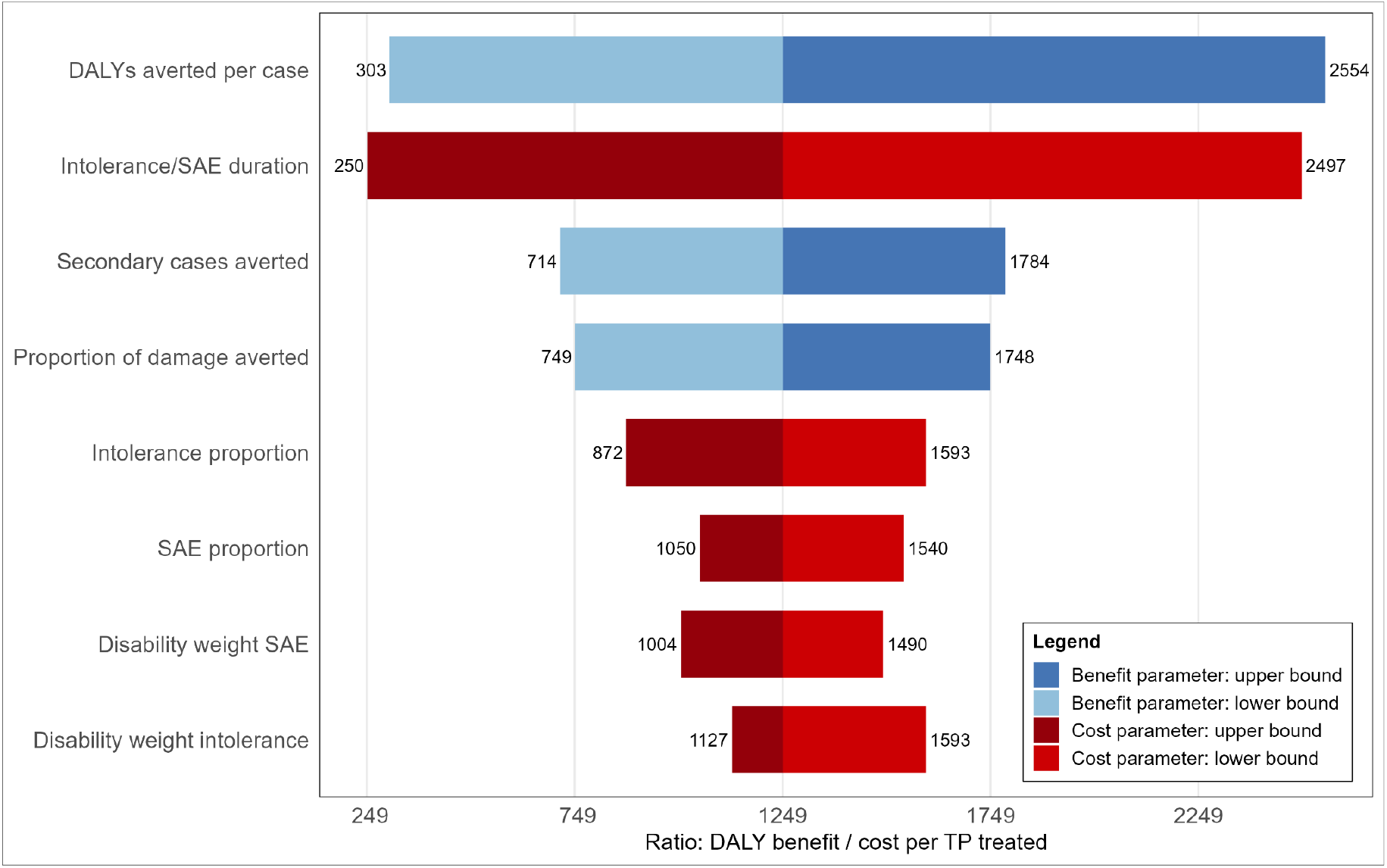
Ratio in Disability-Adjusted Life-Years between appropriate (blue) and overtreatment (red) Figure shows the point ratio in DALYs averted by appropriate treatment (blue bars) and incurred by treatment (red bars). Width of bars shows change in ratio across range for each component (see Table 2) change in DALYs averted (blue tornado) and incurred (red tornado) by varying each component across the specified range. Abbreviations: DALYs - disability adjusted life years; SAE - severe adverse events.

Figure 3 and Supplementary Figure 6A highlight that uncertainty in ranges for estimated DALYs averted per TP treatment was the largest source of variation in the benefit of treatment. For the harm of treatment, the largest driver was the duration of the serious adverse events or intolerance (Figure 3 and Supplementary Figure 6B).

In a hypothetical population of 100,000 individuals with 0.5% prevalence of culture-confirmed pulmonary TB, initiating treatment based on Xpert Ultra averted approximately 60% more DALYs as compared to restricting treatment to culture-confirmed diagnoses only (median 5,977 versus 3,750), with the same ratios for populations with sputum culture-confirmed prevalences of 0.25% and 0.1% (Supplementary Table 12).

### Sensitivity analyses

When using lower baseline sensitivity values of 70%, 60%, and 50%, we found similar trends across the FP:TP ratio (Supplementary Table 7-9 and Supplementary Figure 3-5), as well as the relative DALY benefit of treating based on Xpert Ultra versus restricting to sputum culture-positive individuals only in a population with 0.5% prevalence of culture-confirmed pulmonary TB (Supplementary Table 13-15).

## Discussion

### Main findings

Our results show the limitations of relying solely on culture in correctly identifying individuals who would benefit from TB treatment, leading to false negative diagnoses. In turn, we have consistently underestimated the performance of PCR-based tools, like Xpert Ultra, where the presence of *Mtb* DNA is shown even at low bacterial loads, where culture has struggled. (20) Overall, it appears that following a positive PCR test in community-wide screening, a negative culture is as or more likely to be a false negative as the PCR test is a false positive.

Not all PCR-positive/culture-negative individuals will progress within two years. This most likely reflects the natural history of TB, where recovery in the absence of treatment is well-known, and is more likely when bacterial load is lower. (34,35) However, this does not negate the net benefit of providing treatment, which appears to vastly outweigh the harm. Notably, we already prescribe TB preventive treatment for household contacts, which is effective in this population despite having a much lower risk of developing disease. (36)

Our work implies that treatment could be provided based on PCR results alone, which would bring welcome simplicity in community-wide screening protocols, hopefully reducing the risk of losses along the care cascade, e.g. during the wait for culture results. Given the relatively low bacterial load of community-diagnosed TB (17,37), a shorter regimen could further reduce the potential harms of overtreatment.

Another key finding is that beyond target population and prevalence, specificity (and sensitivity) are also affected by the reference standard used. The ‘true’ specificity of Xpert Ultra tests likely sits in the 99.7-99.9% range, reducing the risk of inappropriate treatment, as observed in practice. (9)

### Limitations

Our work has some limitations. We extended data from a single community study to estimate the proportions of Xpert Ultra trace-positive, culture-negative individuals needing (and assumed to benefit from) treatment, which is a narrow evidence base. However, similar trends were reported during long-term follow-up of individuals with baseline trace-positive results in clinic settings in Uganda and South Africa. (38) Further follow-up studies using similar methods, and evaluating all PCR-positive/culture-negative individuals, would strengthen this assumption. However, the overwhelming benefit over harm found here supports our qualitative findings on the benefits of treating based on the PCR test, which likely stand.

Our baseline sensitivity of 82.5% is pooled from prevalence studies using Xpert Ultra with trace. While perhaps lower with declining prevalence, our sensitivity analyses show our results are robust.

We also highlight the potential value of the trace result in Xpert Ultra in identifying individuals who might benefit from treatment. New sampling approaches, such as tongue-swab-based tests, appear to have reduced yield among individuals with trace-like low bacterial loads (39), reducing this potential benefit of using PCR technology. However, this could be addressed by potential mixed sample approaches (e.g. take sputum where possible, and tongue swab if not) to soften that limitation.

We did not explicitly include some key potential negative impacts of treatment, e.g. catastrophic costs or stigma, mostly because these have not yet been translated to summary health measures (i.e., DALYs). However, the impact on our findings is likely limited, as averting these impacts was also omitted when considering the benefit of treatment, in particular in terms of preventing new episodes of TB. Overall, our current understanding of the net benefit or harm of treatment is poor and should be improved in this era of person-centred care.

Another limitation is that our findings are focussed on treatment initiation after community-wide screening, whereas different considerations may apply in the context of trials for new TB treatment or vaccine trials, in particular given the relative consequences consequence of false-positive and false-negative test results in non-inferiority trials. (40)

### Implications

Broadly, our work highlights the limitations of the current reference standard of *Mtb* growth on sputum culture, likely due to the death of bacteria prior to incubation. The resulting FN test results might have significantly misled our estimates of test performance (8), TB burden, and where we can implement community-wide screening, as well as who should receive treatment. When evaluating diagnostic tools, we should instead consider what or who we are trying to identify? Is it the presence of viable *Mtb*, the presence of *Mtb* DNA, the clinical need for treatment now, or individuals who likely benefit from treatment in the near future? We propose the latter, and pose that a PCR test, which detects *Mtb* DNA, could do a better job than sputum culture at identifying those for whom treatment likely provides benefit, rather than harm.

### Conclusion

Withholding treatment after a positive bacteriological test in community-wide screening is likely more harmful than any harm from overtreatment, supporting expanded coverage of simplified community-wide screening approaches.

## Data Availability

All data produced in the present study are available upon reasonable request to the authors

## Acknowledgements

RMGJH conceived the study. LDV and RMGJH led the analyses and wrote the first draft, with input from AS and KH. All authors provided input on analysis and draft text, and reviewed the final version before submission.

