## Supplementary materials for "Do no harm - re-evaluating the risks of overtreatment in community-wide tuberculosis screening"

Supplementary Material for *Do no harm - re-evaluating the risks of overtreatment in community tuberculosis screening*

Data and scripts available on https://github.com/lshtm-tbmg/Donoharm

### Supplementary Material 1 – Prevalence threshold and coverage of community screening

#### A. Estimating country tuberculosis prevalence in 2023

We aimed to estimate the tuberculosis burden targeted at different tuberculosis screening prevalence thresholds among individuals ≥15 years in 2023. The adult population was chosen because WHO's 0.5% screening threshold is derived from the ACT3 trial [1], which involved participants aged 15 years and above.

**A1. Methods**

A simple national-level prevalence-to-incidence ratio, based on WHO estimates from 2000–2014, was applied to each 2023 WHO country-level incidence estimate to approximate the prevalence of adult TB.

1) Country-level incidence of individuals ≥15 years was retrieved from the GTB 2025 report estimates [2].
2) The average country-level prevalence-to-incidence ratio ("previnc") from 2000-2014 was calculated from WHO estimates.
3) The previnc ratio was applied to the incidence estimate to approximate the prevalence in 2023, to replace 'disease duration', since this is not an universal estimate (e.g. country difference due to access to tuberculosis care):

   a. *Prevalence=incidence*disease duration*
   b. *Prevalence=incidence*previnc*
   c. *Estimated country level tuberculosis prevalence in 2023 = WHO 2023 incidence estimate * average WHO prevalence to incidence estimate from 2000-2014*

<u>Limitations</u>

- In 2015, the WHO discontinued reporting estimated prevalence, as it was no longer included as a metric in the end TB strategy.
- It is unclear whether asymptomatic tuberculosis was included in previous WHO estimations, and therefore how this may have affected the prevalence-to-incidence ratio.
- The prevalence-to-incidence ratio was based on incidence and prevalence estimates for all ages, not restricted to individuals ≥15 years.
- WHO estimates of all tuberculosis were used and may have included extrapulmonary and/or clinically diagnosed TB.

### A2. Results

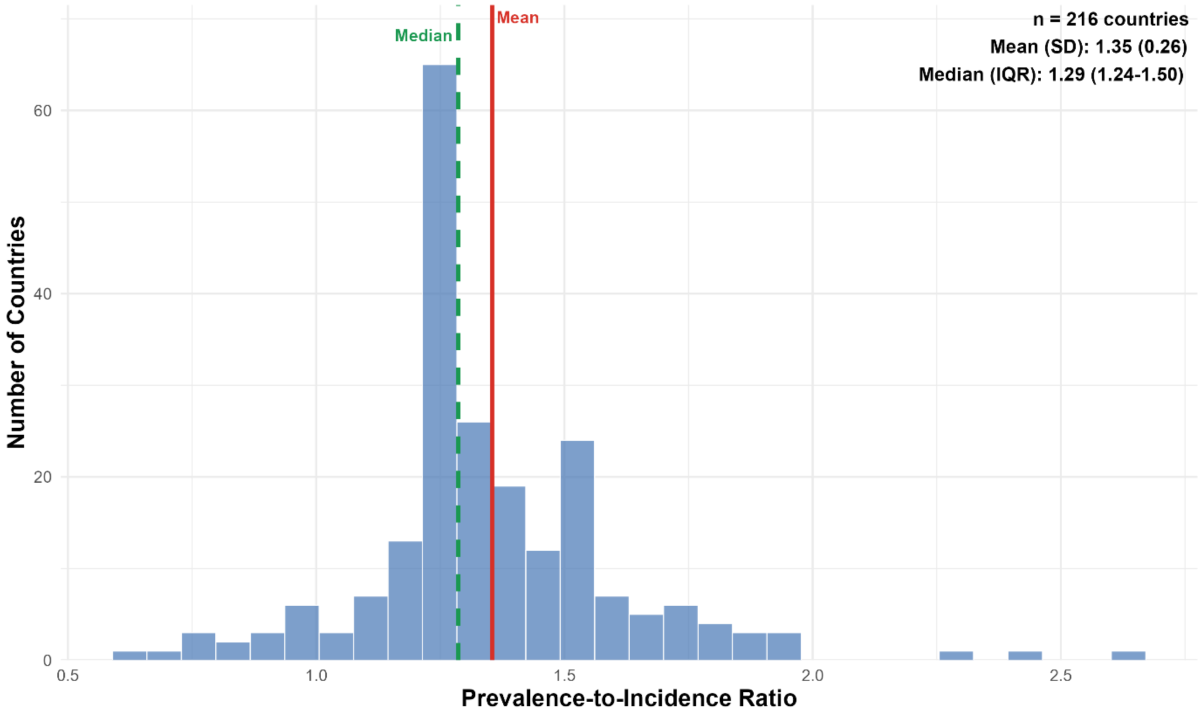

**Supplementary Figure 1.** Distribution of the average country-level prevalence-to-incidence ratio based on WHO estimates from 2000–2014

**Supplementary Table 1.** Adult tuberculosis burden targeted by tuberculosis prevalence

| Threshold tuberculosis prevalence | Proportion global TB burden targeted *N (%)* | Countries included (out of 212) *N (%)* |
|---|---|---|
| ≥0.5% | 6,048,254 (42) | 27 (13) |
| ≥0.25% | 12,102,127 (84) | 53 (25) |
| ≥0.1% | 12,824,017 (89) | 95 (45) |
| ≥0.05% | 14,253,628 (99) | 122 (58) |

**Supplementary Table 2.** Top 31 contributors to global tuberculosis burden among individuals ≥15 years

| ISO | Prevalence (%) (low-high) | Prevalence (1000s) (low-high) | Country contribution (%) | Cumulative contribution (%) | Cumulative contribution (1000s) |
|---|---|---|---|---|---|
| IND | 0.37 (0.3 - 0.44) | 4,020 (3,294 - 4,763) | 27.91 | 27.91 | 4,020 |
| IDN | 0.71 (0.63 - 0.8) | 1,505 (1,320 - 1,691) | 10.44 | 38.35 | 5,525 |
| PHL | 1.46 (0.45 - 2.48) | 1,199 (371 - 2,030) | 8.32 | 46.68 | 6,724 |
| CHN | 0.09 (0.07 - 0.1) | 1,037 (862 - 1,210) | 7.20 | 53.87 | 7,760 |
| PAK | 0.55 (0.35 - 0.76) | 861 (542 - 1,181) | 5.98 | 59.85 | 8,622 |
| BGD | 0.54 (0.37 - 0.72) | 666 (452 - 881) | 4.62 | 64.48 | 9,288 |
| COD | 0.78 (0.43 - 1.14) | 446 (243 - 648) | 3.09 | 67.57 | 9,733 |
| NGA | 0.32 (0.18 - 0.46) | 431 (246 - 617) | 2.99 | 70.56 | 10,165 |
| MMR | 0.97 (0.47 - 1.47) | 397 (194 - 601) | 2.76 | 73.32 | 10,562 |
| VNM | 0.36 (0.2 - 0.52) | 278 (157 - 398) | 1.93 | 75.25 | 10,840 |
| ZAF | 0.45 (0.24 - 0.66) | 211 (113 - 307) | 1.46 | 76.71 | 11,050 |
| ETH | 0.2 (0.12 - 0.28) | 158 (94 - 222) | 1.09 | 77.81 | 11,208 |
| TZA | 0.4 (0.05 - 0.76) | 153 (18 - 290) | 1.06 | 78.87 | 11,361 |
| PRK | 0.71 (0.6 - 0.82) | 153 (128 - 176) | 1.06 | 79.93 | 11,514 |
| THA | 0.25 (0.17 - 0.33) | 151 (102 - 201) | 1.05 | 80.98 | 11,665 |
| AGO | 0.71 (0.34 - 1.09) | 145 (69 - 223) | 1.01 | 81.99 | 11,811 |
| BRA | 0.07 (0.06 - 0.08) | 119 (100 - 139) | 0.83 | 82.82 | 11,930 |
| MOZ | 0.63 (0.31 - 0.95) | 118 (59 - 177) | 0.82 | 83.64 | 12,047 |
| MDG | 0.61 (0.33 - 0.88) | 115 (63 - 167) | 0.80 | 84.43 | 12,162 |
| AFG | 0.48 (0.24 - 0.72) | 112 (56 - 168) | 0.78 | 85.21 | 12,274 |
| KHM | 0.86 (0.42 - 1.3) | 105 (51 - 159) | 0.73 | 85.94 | 12,380 |
| KEN | 0.28 (0.13 - 0.43) | 97 (44 - 149) | 0.67 | 86.61 | 12,476 |
| NPL | 0.4 (0.19 - 0.62) | 86 (39 - 132) | 0.59 | 87.21 | 12,562 |
| UGA | 0.29 (0.14 - 0.43) | 78 (38 - 118) | 0.54 | 87.75 | 12,640 |
| RUS | 0.06 (0.03 - 0.1) | 76 (36 - 117) | 0.53 | 88.28 | 12,716 |
| PER | 0.29 (0.17 - 0.41) | 73 (43 - 104) | 0.51 | 88.79 | 12,789 |
| GHA | 0.32 (0.07 - 0.58) | 70 (16 - 124) | 0.49 | 89.27 | 12,860 |
| SOM | 0.64 (0.31 - 0.97) | 63 (31 - 95) | 0.44 | 89.71 | 12,922 |
| UKR | 0.16 (0.08 - 0.24) | 53 (27 - 79) | 0.37 | 90.08 | 12,975 |
| MYS | 0.19 (0.13 - 0.25) | 52 (36 - 67) | 0.36 | 90.44 | 13,027 |
| PNG | 0.75 (0.55 - 0.94) | 51 (38 - 65) | 0.36 | 90.79 | 13,078 |

**B. Comparison 2023 country-level adult tuberculosis prevalence estimates to recorded prevalence survey bacteriologically-confirmed tuberculosis estimate**

**B1. Methods**

We compared the prevalence survey results of bacteriologically-confirmed adult (≥15 years) [3] tuberculosis prevalence to our adult (≥15 years) tuberculosis prevalence estimates for 2023. The estimates were not matched to the exact year of each survey because the GTB 2024 report provides age-segregated data only for 2023, not for earlier years. For countries with more than one prevalence survey, only the most recent survey was included (31 countries in total).

**B2. Results**

**Supplementary figure 2.** Comparison of tuberculosis prevalence estimates from the study and national prevalence survey estimates

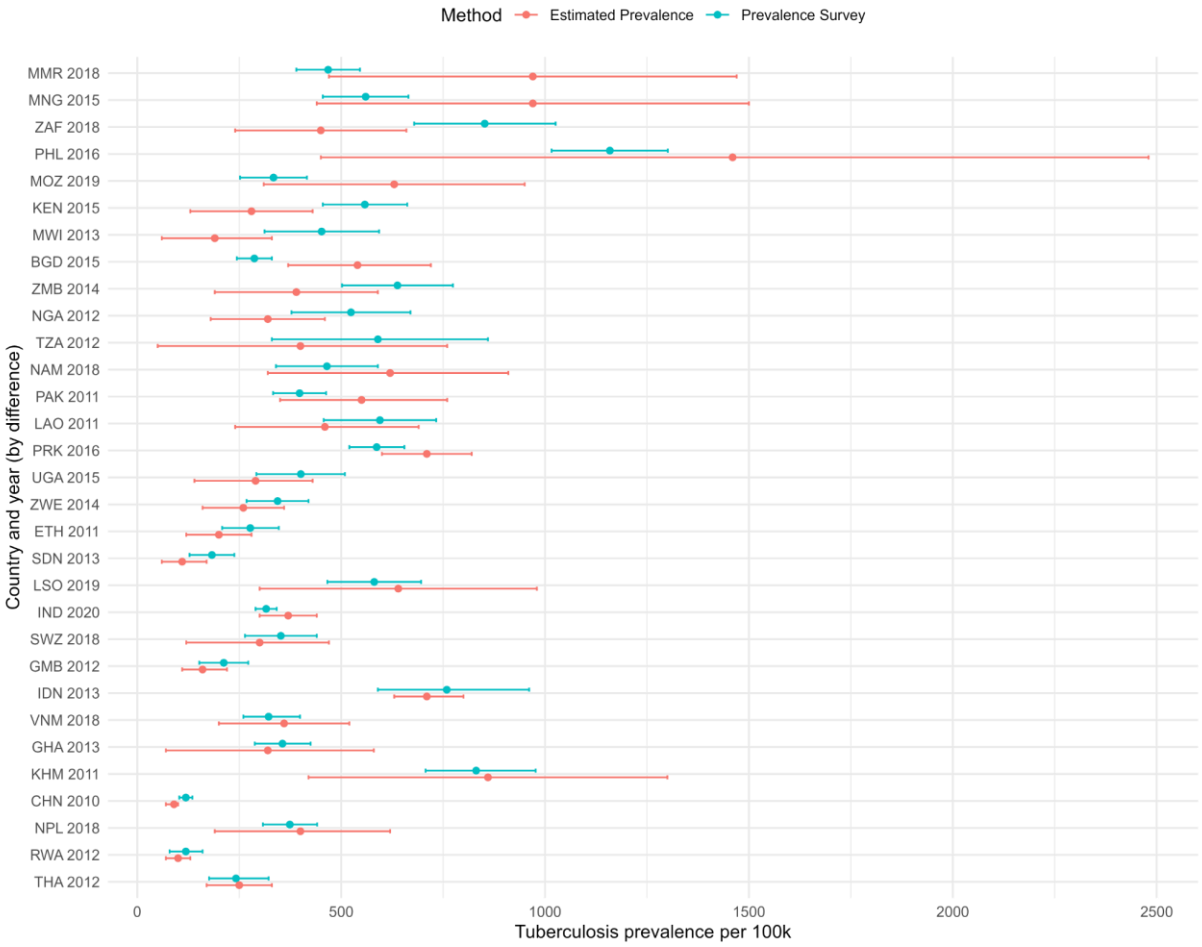

**Supplementary Table 3.** Difference in tuberculosis prevalence estimates from the study and national prevalence survey estimates

| | Prevalence survey estimate **lower than** estimated prevalence | Prevalence survey estimate **bigger than** estimated prevalence |
|---|---|---|
| Difference >50/100k | 10 | 13 |
| Difference >100/100k | 8 | 8 |
| Difference >200/100k | 5 | 5 |

### Supplementary Material 2 - Test characteristics by reference standard

We aimed to estimate, against different reference standard scenarios, the ratio of individuals with an initial positive Xpert Ultra result from community-wide screening in low-prevalence settings that would benefit from treatment (appropriate treatment) to those who would not (overtreatment).

#### A. Data

**Supplementary Table 4.** Xpert Ultra and culture data extracted from Supplementary Table S5 Kendall *et al.* 2021 [4]

| Xpert Ultra result | Culture positive | Culture negative | Total culture positive |
|---|---|---|---|
| Ultra positive, trace | 8 (23%) | 48 (81%) | 56 (60%) |
| Ultra positive, no trace | 27 (77%) | 11 (19%) | 38 (40%) |
| *All Ultra positive* | *35 (100%)* | *59 (100%)* | *94 (100%)* |

**Supplementary Table 5.** Treatment recommendation data among individuals with trace-positive sputum extracted from Sung *et al.* 2025 [5]

| Timepoint | Total evaluated | Recommended for treatment[1] | Microbiological confirmation[2] | Culture positive | Positive 2nd Ultra, urine LAM (negative culture) | Clinically diagnosed[3] | Data informing shift FP to TP with extended reference standards |
|---|---|---|---|---|---|---|---|
| Baseline | 128 | 45 (35%) | 36/45 (80%) | 27/36 (75%) | 9/36 (25%) | 9/45 (20%) | **18/101 (18%)** of trace-positive but culture-negative were recommended for treatment at baseline |
| Follow-up[4] | 75 | 19 (25%) | 15/19 (79%) | — | — | 4/19 (21%) | **19/75 (25%)** of trace-positives, not recommended treatment at baseline were recommended treatment during follow-up |

Footnotes:
Abbreviations: FP – false positive; LAM – lipoarabinomannan; TP – true positive

[1]Excludes cases with presumed false positive microbiological results that were not classified as 'recommended for treatment' (2 at baseline, 4 during follow-up).
[2]Microbiological confirmation includes positive results from liquid and/or solid culture, a second sputum Ultra, or urine LAM test (if HIV positive). See Figure 1 Sung *et al.* 2025 [5] for the evaluation study procedures.
[3]Clinically diagnosed cases lacked microbiological confirmation.
[4]Follow-up group includes individuals without a treatment recommendation at baseline, followed for ≥3 months and up to 2 years; 8 lost to follow-up.

### B. Methods

**Supplementary Table 6.** Equations to estimate the number of true and false positives per reference standard scenario

| Scenario | Individuals assumed to benefit from treatment | Total number true positives individuals | Total number of false positive individuals |
|---|---|---|---|
| Ultra VS culture (all) | 1. Ultra-pos, culture-pos | 1. Population * culture-positive prevalence * sensitivity[1] | 1. (Population – (population * culture-positive prevalence)) * (1 – specificity[2]) |
| Ultra VS culture (all) + baseline Rx evaluation (among Ultra positive only)[c] | 1. Ultra-pos, culture-pos<br>2. 1+ Ultra-pos, culture-neg with baseline Rx recommendation | 1. Population * culture-positive prevalence * sensitivity[1]<br>2. TP in step 1 + (FP in step 1 * 0.18) | 1. (Population – (population * culture-positive prevalence)) * (1 – specificity[2])<br>2. FP in step 1 – (FP in step 1 * 0.18) |
| Ultra VS culture (all) + baseline & follow-up Rx evaluation (among Ultra positive only)[3] | 1. Ultra-pos, culture-pos<br>2. 1 + Ultra-pos, culture-neg with baseline Rx recommendation<br>3. 1 + 2 + Ultra-pos, culture-neg without baseline Rx recommendation but recommended during follow-up (up to 2 years) | 1. Population * culture-positive prevalence * sensitivity[1]<br>2. TP in step 1 + (FP in step 1 * 0.18)<br>3. TP in step 1 + (FP in step 1 * 0.18) + (FP in step 2 * 0.25) | 1. (Population – (population * culture-positive prevalence)) * (1 – specificity[2])<br>2. FP in step 1 – (FP in step 1 * 0.18)<br>3. FP in step 2 – (FP in step 2 * 0.25) |

Footnotes:
Abbreviations: FP – false positives; neg – negative; pos – positive; Rx – treatment; TP – true positives

[1]Sensitivity (Xpert Ultra vs culture) values: primary analysis with an informed sensitivity of 82.5% (pooled sensitivity Xpert Ultra with trace as positive from prevalence surveys [6]) and sensitivity analyses with sensitivity values of 70% (WHO's optimal sensitivity value in combination with a 98% specificity in a high-specificity screening test scenario [7]), 60% (WHO's minimal sensitivity value in combination with a 98% specificity in a high-specificity screening test scenario [7]), 50% (uninformed).
[2]Specificity (Xpert Ultra vs culture) values = using the estimated linear regression between prevalence and specificity of Xpert Ultra with trace as positive found among unenriched community-wide screening populations and screen-positive prevalence survey adult populations [8].
[3]Proportions of FP shifting to TP based on data Sung *et al.* 2025 [5], as reported in Supplementary Table 5 above.

Note: The proportion additionally recommended for treatment at baseline and during follow-up among ultra-positive, culture-negative individuals may be a conservative estimate. These proportions were derived from data of trace-positives only from a community screening in Uganda [5] but were applied to all Xpert Ultra–positive, culture-negative cases. It may be assumed that the proportions would have been higher among individuals with Xpert Ultra results above the trace level. However, based on Supplementary Table 4, among all Ultra-positive but culture-negative cases, only 19% had results above the trace level, while the remaining 81% were trace-positive in a Ugandan community screening [4].

**C. Results**

**Supplementary Table 7.** Screening test performance with extended reference standard assuming an Xpert Ultra sensitivity of 70%, among a hypothetical population of 100,000 individuals

| Culture+ Prevalence (%) | Scenario | FP | TP | TN | FN | prev (%) | spec (%) | sens (%) | FP per TP | PPV (%) | NPV (%) | NNS | NNH |
|---|---|---|---|---|---|---|---|---|---|---|---|---|---|
| 0.5 | Ultra VS culture | 498 | 350 | 99,002 | 150 | 0.500 | 99.5 | 70.0 | 1.42 | 41.3 | 99.8 | 286 | 201 |
| | Ultra VS culture + baseline Rx evaluation (among Ultra positive only) | 408 | 440 | 99,002 | 150 | 0.590 | 99.6 | 74.6 | 0.93 | 51.9 | 99.8 | 227 | 245 |
| | Ultra VS culture + baseline & follow-up Rx evaluation (among Ultra positive only) | 306 | 542 | 99,002 | 150 | 0.692 | 99.7 | 78.3 | 0.56 | 63.9 | 99.8 | 185 | 327 |
| 0.25 | Ultra VS culture | 252 | 175 | 99,498 | 75 | 0.250 | 99.7 | 70.0 | 1.44 | 41.0 | 99.9 | 571 | 397 |
| | Ultra VS culture + baseline Rx evaluation (among Ultra positive only) | 207 | 220 | 99,498 | 75 | 0.295 | 99.8 | 74.6 | 0.94 | 51.5 | 99.9 | 455 | 483 |
| | Ultra VS culture + baseline & follow-up Rx evaluation (among Ultra positive only) | 155 | 272 | 99,498 | 75 | 0.347 | 99.8 | 78.4 | 0.57 | 63.7 | 99.9 | 368 | 645 |
| 0.1 | Ultra VS culture | 104 | 70 | 99,796 | 30 | 0.100 | 99.9 | 70.0 | 1.49 | 40.2 | 100.0 | 1,429 | 962 |
| | Ultra VS culture + baseline Rx evaluation (among Ultra positive only) | 85 | 89 | 99,796 | 30 | 0.119 | 99.9 | 74.8 | 0.96 | 51.1 | 100.0 | 1,124 | 1,176 |
| | Ultra VS culture + baseline & follow-up Rx evaluation (among Ultra positive only) | 64 | 110 | 99,796 | 30 | 0.140 | 99.9 | 78.6 | 0.58 | 63.2 | 100.0 | 909 | 1,562 |

Abbreviations: FP – false positives; NNH – number needed to harm; NNS – number needed to screen; NPV – negative predictive value; PPV – positive predictive value; prev – prevalence; Rx – treatment; sens – sensitivity; spec – specificity; TN – true negatives; TP – true positives.

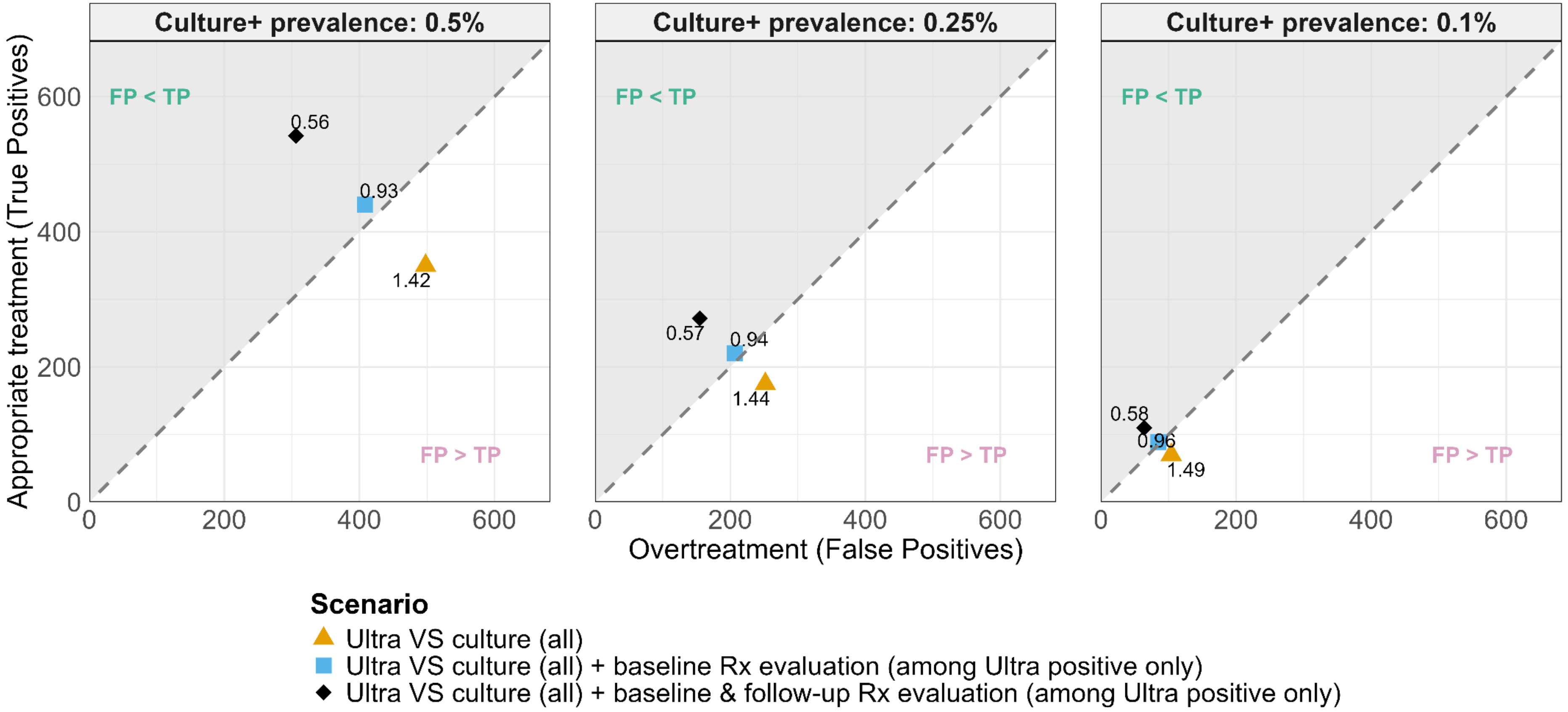

**Supplementary Figure 3.** Balance between overtreatment and appropriate treatment assuming an Xpert Ultra sensitivity of 70%, among a hypothetical population of 100,000 individuals

**Supplementary Table 8.** Screening test performance with extended reference standard assuming an Xpert Ultra sensitivity of 60%, among a hypothetical population of 100,000 individuals

| Culture+ Prevalence (%) | Scenario | FP | TP | TN | FN | prev (%) | spec (%) | sens (%) | FP per TP | PPV (%) | NPV (%) | NNS | NNH |
|---|---|---|---|---|---|---|---|---|---|---|---|---|---|
| 0.5 | Ultra VS culture | 498 | 300 | 99,002 | 200 | 0.500 | 99.5 | 60.0 | 1.66 | 37.6 | 99.8 | 333 | 201 |
| | Ultra VS culture + baseline Rx evaluation (among Ultra positive only) | 408 | 390 | 99,002 | 200 | 0.590 | 99.6 | 66.1 | 1.05 | 48.9 | 99.8 | 256 | 245 |
| | Ultra VS culture + baseline & follow-up Rx evaluation (among Ultra positive only) | 306 | 492 | 99,002 | 200 | 0.692 | 99.7 | 71.1 | 0.62 | 61.7 | 99.8 | 203 | 327 |
| 0.25 | Ultra VS culture | 252 | 150 | 99,498 | 100 | 0.250 | 99.7 | 60.0 | 1.68 | 37.3 | 99.9 | 667 | 397 |
| | Ultra VS culture + baseline Rx evaluation (among Ultra positive only) | 207 | 195 | 99,498 | 100 | 0.295 | 99.8 | 66.1 | 1.06 | 48.5 | 99.9 | 513 | 483 |
| | Ultra VS culture + baseline & follow-up Rx evaluation (among Ultra positive only) | 155 | 247 | 99,498 | 100 | 0.347 | 99.8 | 71.2 | 0.63 | 61.4 | 99.9 | 405 | 645 |
| 0.1 | Ultra VS culture | 104 | 60 | 99,796 | 40 | 0.100 | 99.9 | 60.0 | 1.73 | 36.6 | 100.0 | 1,667 | 962 |
| | Ultra VS culture + baseline Rx evaluation (among Ultra positive only) | 85 | 79 | 99,796 | 40 | 0.119 | 99.9 | 66.4 | 1.08 | 48.2 | 100.0 | 1,266 | 1,176 |
| | Ultra VS culture + baseline & follow-up Rx evaluation (among Ultra positive only) | 64 | 100 | 99,796 | 40 | 0.140 | 99.9 | 71.4 | 0.64 | 61.0 | 100.0 | 1,000 | 1,562 |

Abbreviations: FN – false negative; FP – false positives; NNH – number needed to harm; NNS – number needed to screen; NPV – negative predictive value; PPV – positive predictive value; prev – prevalence; Rx – treatment; sens – sensitivity; spec – specificity; TN – true negatives; TP – true positives.

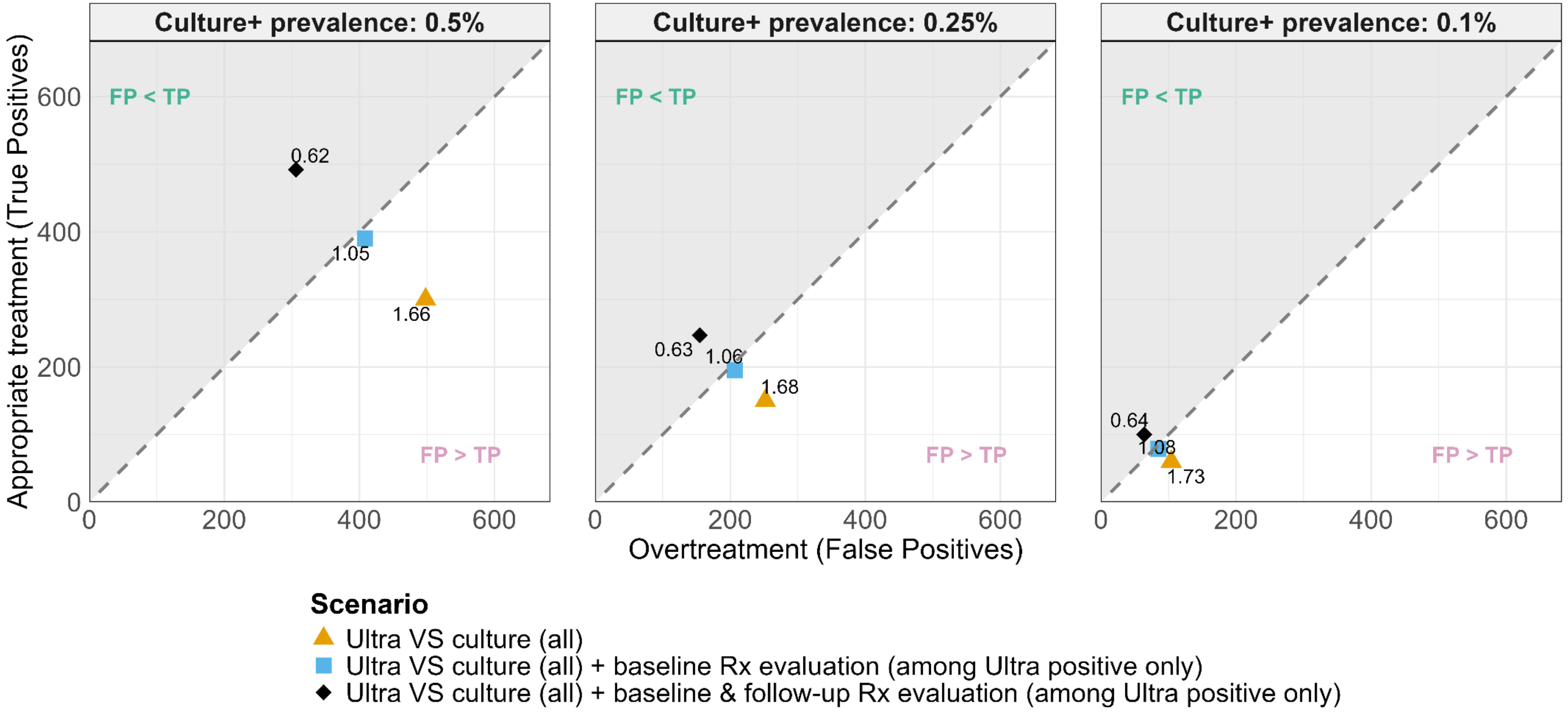

**Supplementary Figure 4.** Balance between overtreatment and appropriate treatment assuming an Xpert Ultra sensitivity of 60%, among a hypothetical population of 100,000 individuals

**Supplementary Table 9.** Screening test performance with extended reference standard assuming an Xpert Ultra sensitivity of 50%, among a hypothetical population of 100,000 individuals

| Culture+ Prevalence (%) | Scenario | FP | TP | TN | FN | prev (%) | spec (%) | sens (%) | FP per TP | PPV (%) | NPV (%) | NNS | NNH |
|---|---|---|---|---|---|---|---|---|---|---|---|---|---|
| 0.5 | Ultra VS culture | 498 | 250 | 99,002 | 250 | 0.500 | 99.5 | 50.0 | 1.99 | 33.4 | 99.7 | 400 | 201 |
| | Ultra VS culture + baseline Rx evaluation (among Ultra positive only) | 408 | 340 | 99,002 | 250 | 0.590 | 99.6 | 57.6 | 1.20 | 45.5 | 99.7 | 294 | 245 |
| | Ultra VS culture + baseline & follow-up Rx evaluation (among Ultra positive only) | 306 | 442 | 99,002 | 250 | 0.692 | 99.7 | 63.9 | 0.69 | 59.1 | 99.7 | 226 | 327 |
| 0.25 | Ultra VS culture | 252 | 125 | 99,498 | 125 | 0.250 | 99.7 | 50.0 | 2.02 | 33.2 | 99.9 | 800 | 397 |
| | Ultra VS culture + baseline Rx evaluation (among Ultra positive only) | 207 | 170 | 99,498 | 125 | 0.295 | 99.8 | 57.6 | 1.22 | 45.1 | 99.9 | 588 | 483 |
| | Ultra VS culture + baseline & follow-up Rx evaluation (among Ultra positive only) | 155 | 222 | 99,498 | 125 | 0.347 | 99.8 | 64.0 | 0.70 | 58.9 | 99.9 | 450 | 645 |
| 0.1 | Ultra VS culture | 104 | 50 | 99,796 | 50 | 0.100 | 99.9 | 50.0 | 2.08 | 32.5 | 99.9 | 2,000 | 962 |
| | Ultra VS culture + baseline Rx evaluation (among Ultra positive only) | 85 | 69 | 99,796 | 50 | 0.119 | 99.9 | 58.0 | 1.23 | 44.8 | 99.9 | 1,449 | 1,176 |
| | Ultra VS culture + baseline & follow-up Rx evaluation (among Ultra positive only) | 64 | 90 | 99,796 | 50 | 0.140 | 99.9 | 64.3 | 0.71 | 58.4 | 99.9 | 1,111 | 1,562 |

Abbreviations: FN – false negative; FP – false positives; NNH – number needed to harm; NNS – number needed to screen; NPV – negative predictive value; PPV – positive predictive value; prev – prevalence; Rx – treatment; sens – sensitivity; spec – specificity; TN – true negatives; TP – true positives.

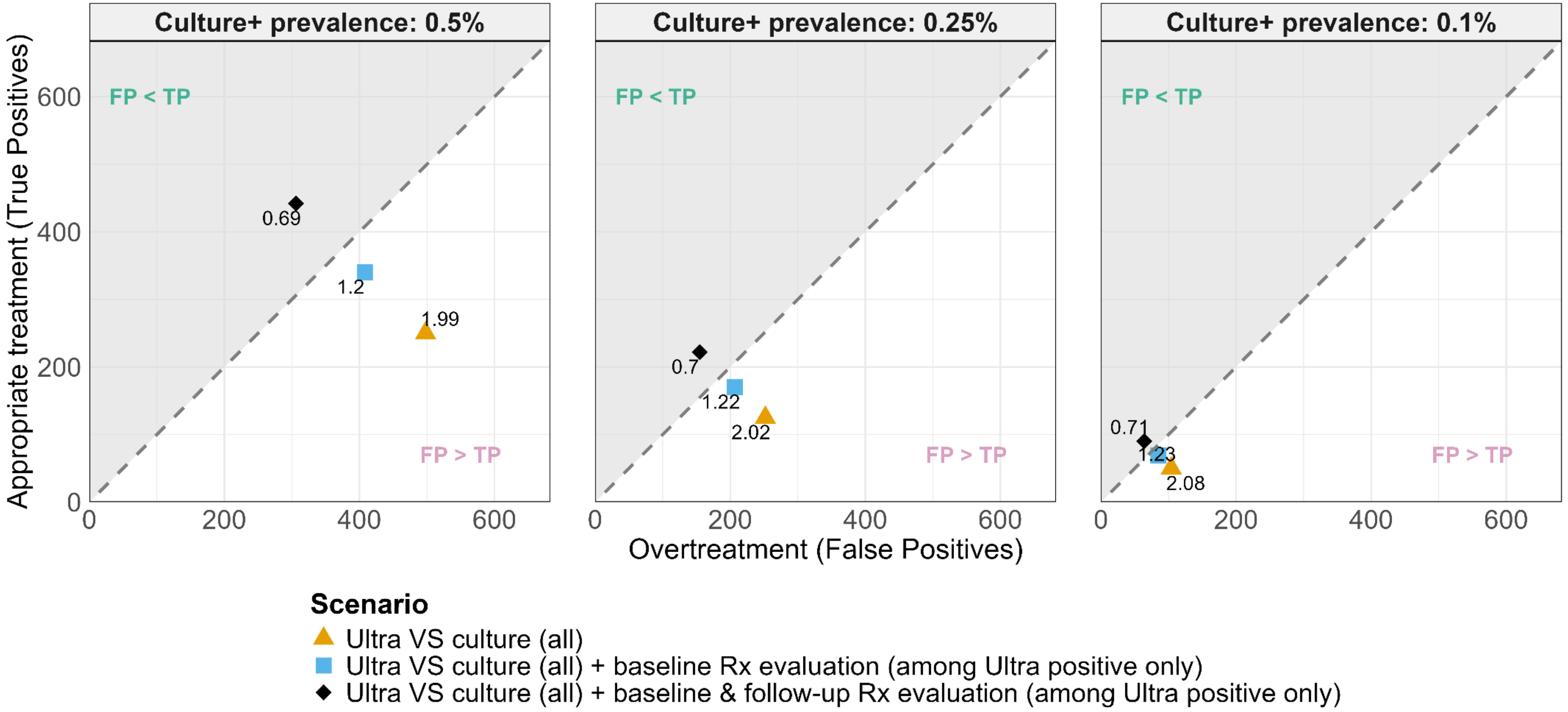

**Supplementary Figure 5.** Balance between overtreatment and appropriate treatment assuming an Xpert Ultra sensitivity of 50%, among a hypothetical population of 100,000 individuals

### Supplementary Material 3 – Harm vs Benefit

#### A. Methods

**Supplementary Table 10.** Definitions used to quantify health benefit and harm

| Measure | Definition | Equation | Assumptions |
|---|---|---|---|
| **General DALY definitions** | | | |
| DALY | Years of life lost (YLL) plus years lived with disability (YLD) | DALY = YLL + YLD | — |
| Individual-level DALY | DALY for a single individual | (standard life expectancy − age at death) + (disability weight * duration [years]) | — |
| **Study definitions** | | | |
| Benefit | Total DALYs averted per individual with infectious TB disease treated | (DALYs averted per case + (secondary cases averted * DALYs averted per case)) * proportion damage averted | — |
| Cost | Total DALYs per individual receiving anti-TB treatment | duration [years] * ((risk SAE × disability weight SAE) + (risk intolerance * disability weight intolerance)) | No years of life lost assumed from treatment |
| Ratio DALYs benefit/cost | Benefit-to-cost ratio per individual with infectious TB disease receiving anti-TB treatment | benefit ÷ cost | — |

Abbreviations: DALYs – disability adjusted life years; SAE – serious adverse events; YLD – years lived with disability; YLL – Years of life lost

**Supplementary Table 11. DALYs incurred by false positive diagnosis, averted by true positive diagnosis, and incurred by false negative diagnosis**

| Scenario | DALYs incurred by FP | DALYs averted TP | DALYs incurred by FN | Net benefit DALYs | Ratio |
|---|---|---|---|---|---|
| **1.** Ultra VS culture (all) | Number of FP * base case cost | number of TP * (base case benefit – base case cost) | Number of FN * base case benefit | DALYs TP – (DALYs FP + DALYs FN) | Reference |
| **2.** Ultra VS culture (all) + baseline Rx evaluation (among Ultra positive only) | | | | | Total DALYs scenario 2 ÷ total DALYs scenario 1 |
| **3.** Ultra VS culture (all) + baseline & follow-up Rx evaluation (among Ultra positive only) | | | | | Total DALYs scenario 3 ÷ total DALYs scenario 1 |

Abbreviations: DALYs – disability adjusted life years; FN – false negative; FP – false positives; Rx - treatment; TP – true positives

**B. Results**

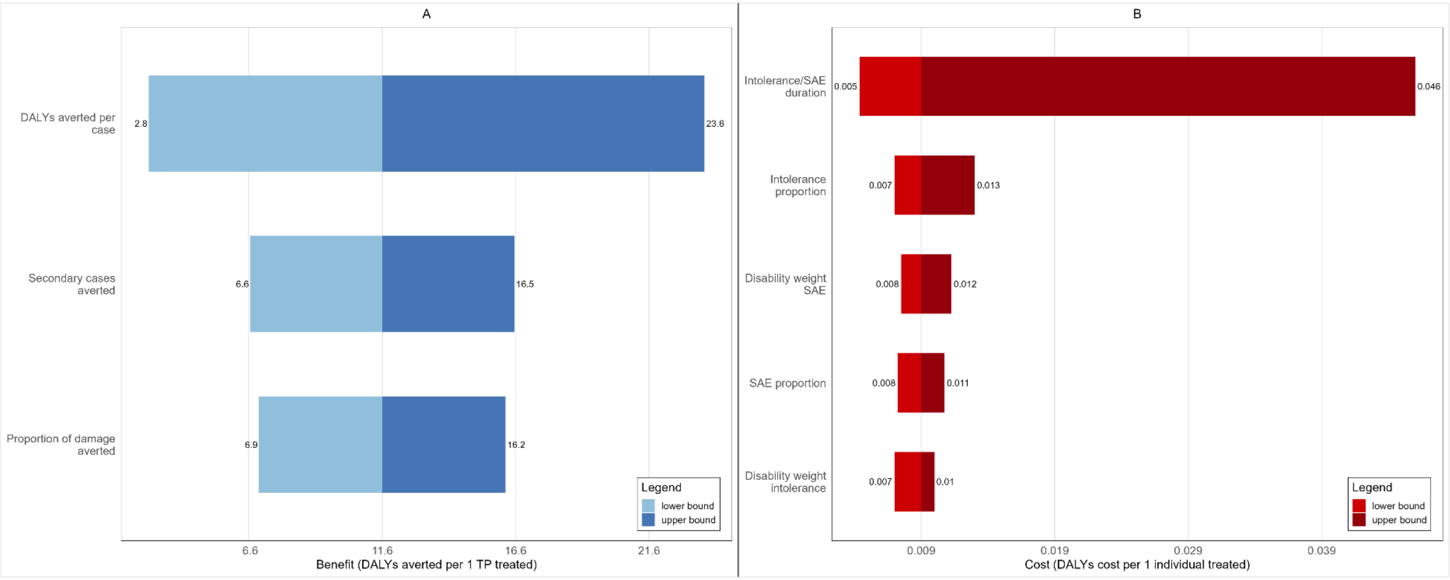

**Supplementary Figure 6.** Disability adjusted life years averted by appropriate treatment (A) and incurred by treatment (B). Base case values: Benefit = 11.6 DALYs; Cost = 0.009 DALYs. Abbreviations: DALYs - disability adjusted life years; SAE - severe adverse events

**Supplementary Table 12.** Balance harm and benefit assuming an Xpert Ultra sensitivity of 82.5%, among a hypothetical population of 100,000 individuals

| Culture-positive prevalence (%) | Scenario | DALYs incurred by FP | DALYs averted by TP | DALYs incurred by FN | Net benefit DALYs | Ratio |
|---|---|---|---|---|---|---|
| 0.5 | Ultra VS culture | 4.48 | 4,775 | 1,021 | 3,750 | Reference |
| | Ultra VS culture + baseline Rx evaluation (among Ultra positive only) | 3.67 | 5,819 | 1,021 | 4,794 | 1.28 |
| | Ultra VS culture + baseline & follow-up Rx evaluation (among Ultra positive only) | 2.75 | 7,001 | 1,021 | 5,977 | 1.59 |
| 0.25 | Ultra VS culture | 2.27 | 2,388 | 510 | 1,876 | Reference |
| | Ultra VS culture + baseline Rx evaluation (among Ultra positive only) | 1.86 | 2,921 | 510 | 2,409 | 1.28 |
| | Ultra VS culture + baseline & follow-up Rx evaluation (among Ultra positive only) | 1.39 | 3,512 | 510 | 3,001 | 1.6 |
| 0.1 | Ultra VS culture | 0.94 | 950 | 209 | 740 | Reference |
| | Ultra VS culture + baseline Rx evaluation (among Ultra positive only) | 0.76 | 1,171 | 209 | 961 | 1.3 |
| | Ultra VS culture + baseline & follow-up Rx evaluation (among Ultra positive only) | 0.58 | 1,426 | 209 | 1,216 | 1.64 |

Abbreviations: DALYs – disability adjusted life years; FN – false negative; FP – false positives; Rx - treatment; TP – true positives

**Supplementary Table 13.** Balance harm and benefit assuming an Xpert Ultra sensitivity of 70%, among a hypothetical population of 100,000 individuals

| Culture-positive prevalence (%) | Scenario | DALYs incurred by FP | DALYs averted by TP | DALYs incurred by FN | Net benefit DALYs | Ratio |
|---|---|---|---|---|---|---|
| 0.5 | Ultra VS culture | 4.48 | 4,057 | 1,740 | 2,313 | Reference |
| | Ultra VS culture + baseline Rx evaluation (among Ultra positive only) | 3.67 | 5,100 | 1,740 | 3,356 | 1.45 |
| | Ultra VS culture + baseline & follow-up Rx evaluation (among Ultra positive only) | 2.75 | 6,282 | 1,740 | 4,539 | 1.96 |
| 0.25 | Ultra VS culture | 2.27 | 2,028 | 870 | 1,156 | Reference |
| | Ultra VS culture + baseline Rx evaluation (among Ultra positive only) | 1.86 | 2,550 | 870 | 1,678 | 1.45 |
| | Ultra VS culture + baseline & follow-up Rx evaluation (among Ultra positive only) | 1.39 | 3,153 | 870 | 2,282 | 1.97 |
| 0.1 | Ultra VS culture | 0.94 | 811 | 348 | 462 | Reference |
| | Ultra VS culture + baseline Rx evaluation (among Ultra positive only) | 0.76 | 1,032 | 348 | 683 | 1.48 |
| | Ultra VS culture + baseline & follow-up Rx evaluation (among Ultra positive only) | 0.58 | 1,275 | 348 | 926 | 2 |

Abbreviations: DALYs – disability adjusted life years; FN – false negative; FP – false positives; Rx - treatment; TP – true positives

**Supplementary Table 14.** Balance harm and benefit assuming an Xpert Ultra sensitivity of 60%, among a hypothetical population of 100,000 individuals

| Culture-positive prevalence (%) | Scenario | DALYs incurred by FP | DALYs averted by TP | DALYs incurred by FN | Net benefit DALYs | Ratio |
|---|---|---|---|---|---|---|
| 0.5 | Ultra VS culture | 4.48 | 3,477 | 2,320 | 1,153 | Reference |
| | Ultra VS culture + baseline Rx evaluation (among Ultra positive only) | 3.67 | 4,520 | 2,320 | 2,196 | 1.9 |
| | Ultra VS culture + baseline & follow-up Rx evaluation (among Ultra positive only) | 2.75 | 5,703 | 2,320 | 3,380 | 2.93 |
| 0.25 | Ultra VS culture | 2.27 | 1,739 | 1,160 | 577 | Reference |
| | Ultra VS culture + baseline Rx evaluation (among Ultra positive only) | 1.86 | 2,260 | 1,160 | 1,098 | 1.9 |
| | Ultra VS culture + baseline & follow-up Rx evaluation (among Ultra positive only) | 1.39 | 2,863 | 1,160 | 1,702 | 2.95 |
| 0.1 | Ultra VS culture | 0.94 | 695 | 464 | 230 | Reference |
| | Ultra VS culture + baseline Rx evaluation (among Ultra positive only) | 0.76 | 916 | 464 | 451 | 1.96 |
| | Ultra VS culture + baseline & follow-up Rx evaluation (among Ultra positive only) | 0.58 | 1,159 | 464 | 694 | 3.02 |

Abbreviations: DALYs – disability adjusted life years; FN – false negative; FP – false positives; Rx - treatment; TP – true positives

**Supplementary Table 15.** Balance harm and benefit assuming an Xpert Ultra sensitivity of 50%, among a hypothetical population of 100,000 individuals

| Culture-positive prevalence (%) | Scenario | DALYs incurred by FP | DALYs averted by TP | DALYs incurred by FN | Net benefit DALYs | Ratio |
|---|---|---|---|---|---|---|
| 0.5 | Ultra VS culture | 4.48 | 2,898 | 2,900 | -6 | Reference |
| | Ultra VS culture + baseline Rx evaluation (among Ultra positive only) | 3.67 | 3,941 | 2,900 | 1,037 | -172.83 |
| | Ultra VS culture + baseline & follow-up Rx evaluation (among Ultra positive only) | 2.75 | 5,123 | 2,900 | 2,220 | -370 |
| 0.25 | Ultra VS culture | 2.27 | 1,449 | 1,450 | -3 | Reference |
| | Ultra VS culture + baseline Rx evaluation (among Ultra positive only) | 1.86 | 1,970 | 1,450 | 518 | -172.67 |
| | Ultra VS culture + baseline & follow-up Rx evaluation (among Ultra positive only) | 1.39 | 2,573 | 1,450 | 1,122 | -374 |
| 0.1 | Ultra VS culture | 0.94 | 580 | 580 | -1 | Reference |
| | Ultra VS culture + baseline Rx evaluation (among Ultra positive only) | 0.76 | 800 | 580 | 219 | -219 |
| | Ultra VS culture + baseline & follow-up Rx evaluation (among Ultra positive only) | 0.58 | 1,043 | 580 | 462 | -462 |

Abbreviations: DALYs – disability adjusted life years; FN – false negative; FP – false positives; Rx - treatment; TP – true positives
